# Disrupted Ca^2+^ homeostasis and immunodeficiency in patients with functional Inositol 1,4,5-trisphosphate receptor subtype 3 defects

**DOI:** 10.1101/2021.05.29.21257775

**Authors:** Julika Neumann, Erika Van Nieuwenhove, Lara E Terry, Frederik Staels, Taylor R Knebel, Kirsten Welkenhuyzen, Mariah R Baker, Margaux Gerbaux, Mathijs Willemsen, John S. Barber, Irina I Serysheva, Liesbeth De Waele, François Vermeulen, Isabelle Meyts, David I Yule, Geert Bultynck, Rik Schrijvers, Stephanie Humblet-Baron, Adrian Liston

**Author notes:** equal contribution (Julika Neumann, Erika Van Nieuwenhove, Lara E Terry, Stephanie Humblet-Baron, Adrian Liston). Clinical correspondence to or.

## Abstract

Calcium signaling is essential for lymphocyte activation, with genetic disruptions resulting in severe immunodeficiency. The inositol 1,4,5-trisphosphate receptor (IP_3_R), formed from homo- or hetero-tetramers of the IP_3_R isoforms 1-3, amplifies lymphocyte signaling by releasing Ca^2+^ from ER stores into the cytosol following antigen-stimulation. While knockout of all 3 IP_3_R isoforms results in immunodeficiency in mice, the seeming redundancy of subunits was thought to explain the absence of IP_3_R mutation as a cause of human immunodeficiency. Here, we identify compound heterozygous variants in *ITPR3* in two unrelated Caucasian patients presenting with combined immunodeficiency, in one case requiring hematopoietic stem cell transplantation. We observed disrupted Calcium homeostasis in patient-derived fibroblasts and immune cells, with abnormal proliferation and activation responses following B and T cell receptor stimulation. Reconstitution of IP_3_R knockout cell lines identified the variants as functional hypomorphs with reduced discrimination between homeostatic and induced states, validating a link between genotype and phenotype. These results demonstrate a functional linkage between defective ER Ca^2+^ channels and immunodeficiency, and identify IP_3_Rs as diagnostic targets for patients with specific inborn errors of immunity.

## Introduction

Spatiotemporally controlled changes in cytosolic Ca^2+^ concentrations serve as a key signaling mediator in a multitude of physiological processes, from neuron excitation through to cellular apoptosis and lymphocyte activation. In lymphocytes, elevation of the cytosolic Ca^2+^ concentration ([Ca^2+^]_cyt_) is a key event following the engagement of the B cell receptor (BCR) or T cell receptor (TCR). In these cells, Ca^2+^ signaling occurs via intracellular efflux and extracellular influx in multiple phases. The first step is mediated by the second messenger inositol 1,4,5-trisphosphate (IP_3_), generated upon activation of phospholipase C (PLC) γ. IP_3_ binds to and opens IP_3_ receptors (IP_3_Rs), thereby releasing Ca^2+^ from endoplasmic reticulum (ER) stores into the cytosol. While this event only slightly increases [Ca^2+^]_cyt_, in the second stage the ER transmembrane protein STIM1 senses lower ER [Ca^2+^], and via a conformational change leads to the opening of plasmalemmal ORAI1 channels. ORAI1 is a calcium-release activated Ca^2+^ channel (CRAC) that mediates influx of extracellular Ca^2+^, a process known as store-operated Ca^2+^ entry. This sustained increase in [Ca^2+^]_cyt_ triggers downstream signaling, notably the NF-κB and calcineurin/NFAT pathways^1^, thereby activating the antigen-stimulated lymphocyte.

Due to the centrality of Ca^2+^ regulation, a large diversity of genetic disorders are associated with disrupted Ca^2+^ pathways^2^. Inadequate Ca^2+^ signaling has long been associated with severe immunodeficiencies, highlighting the importance of functional Ca^2+^ signaling in immune cell activation^3,4^. Genetic defects in extracellular Ca^2+^ influx are formally associated with primary immunodeficiency, with loss of ORAI1 or STIM1 causing severe combined immunodeficiency (SCID)^5,6^. By contrast, the primary ER Ca^2+^ release through IP_3_R channels, although associated with several non-immunological disorders^7,8^, has yet to be linked to primary immunodeficiencies. Variants in *ITPR1, ITPR2* and *ITPR3* have been reported to cause (spino-)cerebellar ataxia^9,10^, anhidrosis^11^, and Charcot-Marie-Tooth disease^12^, respectively. In mice, lymphocyte defects are not observed in individual knockouts, only arising in triple-knockout mice^13–15^. Based on the 60-80% sequence homology amongst the isoforms^16^ and the formation of either homo- or hetero-tetramers, subunit redundancy between subunits may occur in lymphocytes. However, whether lymphocytes are susceptible to genetic variants impairing the primary Ca^2+^ efflux from the ER in humans remained unknown.

Here we identified two patients from unrelated kindreds with immunodeficiency and immune dysregulation that harbor compound heterozygous variants in *ITPR3*. We show that the three variants identified differentially impact channel function and intracellular Calcium homeostasis, with proliferation and activation defects observed in stimulated lymphocytes. Together these results demonstrate that human lymphocytes are sensitive to genetic defects in the primary ER Ca^2+^ release system, with niche-filling *ITPR3* variants altering IP_3_R sensitivity. This establishes genetic defects of IP_3_ receptors as a new class of inborn errors of immunity.

## Results

### *Identification of compound heterozygous* ITPR3 *variants in two immunodeficient kindreds*

To identify the cause of immunodeficiency in two patients without genetic diagnosis, comprehensive clinical, immunological and genetic analysis was performed. Both patients were born to non-consanguineous parents of European descent (**Fig. 1A**). P1 is male patient in his early adolescence who presented with a combined immunodeficiency with severely reduced numbers of B and T cells, requiring hematopoietic stem cell transplantation (HSCT) in the first decade of his life. He developed an EBV-induced leiomyoma after transplantation and showed abnormal tooth eruption and mineralization as well as thin hair since birth. In his early adolescence, P1 presented with peripheral neuropathy and was diagnosed with Charcot-Marie-Tooth. P2 is a male in his 30’s who presented with recurring immune thrombocytopenia (ITP) requiring splenectomy in his late adolescence. He subsequently suffered from autoimmune hemolytic anemia, susceptibility to infections, and enteropathy. Hypogammaglobulinemia and low switched memory B cells evoked diagnosis with CVID and led to treatment with monthly intravenous immunoglobulins (IVIG). Additional clinical details are available in the Supplementary Material (**Supplementary Tables 1-3**). None of the parents showed a clinically apparent immunological phenotype and all were self-reported healthy.

**Figure 1:**
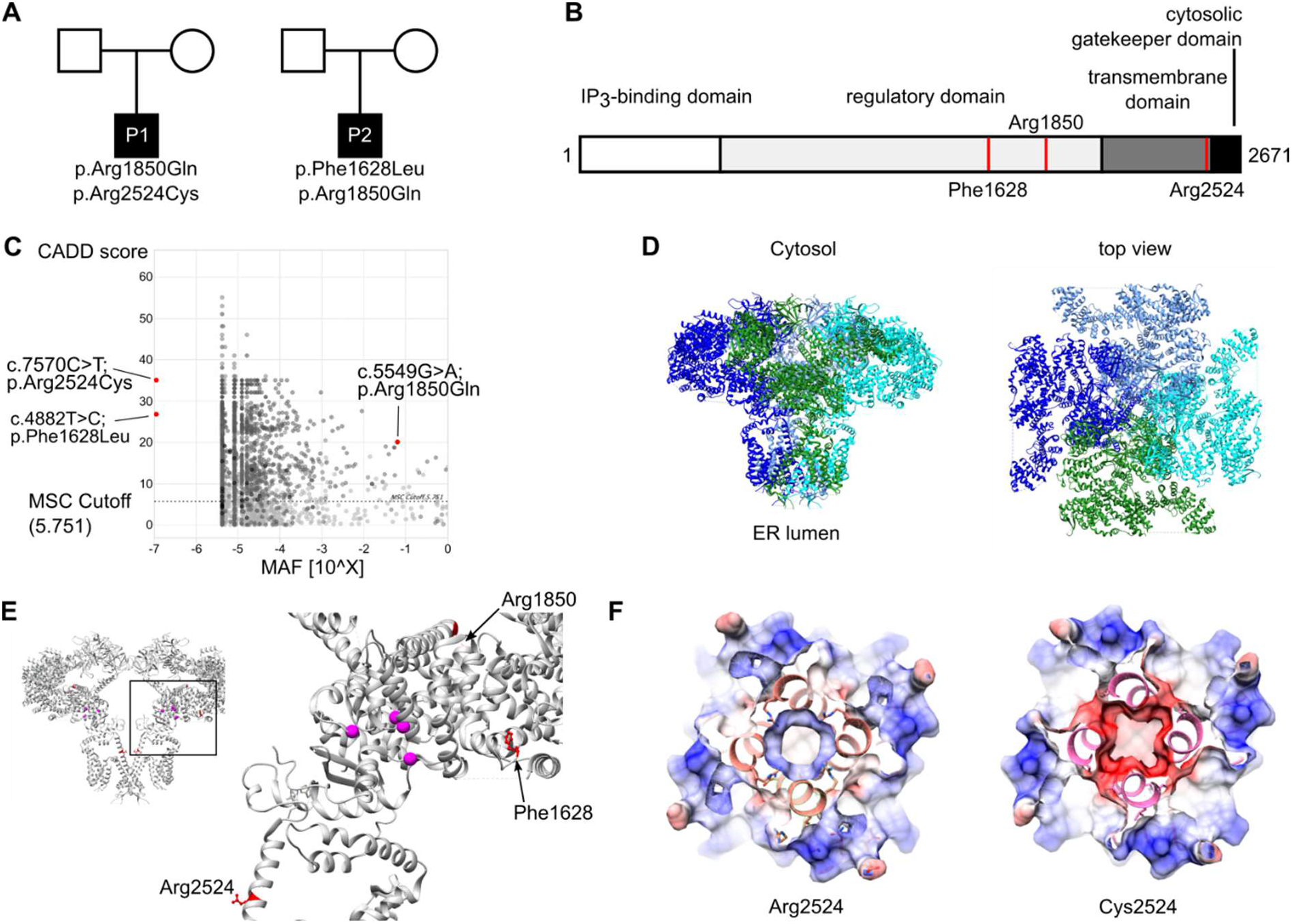
Identification of compound heterozygous variants in *ITPR3* in two kindreds with immunodeficiency or immune dysregulation. **(A)** Pedigrees of two unrelated families, illustrating the genotype of the patients. Specific genotypes of the parents are not disclosed to protect their privacy. **(B)** Domain structure of IP_3_R3 with positions of the variants indicated by red lines. **(C)** PopViz plot showing the mean allelic frequency (MAF) and the Combined Annotation Dependent Deletion (CADD) score for the identified variants in red. The dashed line indicates the gene-specific mutation significance cutoff (MSC). **(D)** Cryo-EM structure of homo-tetrameric IP_3_R3 (6DQJ) depicted as ribbon diagram with different subunits color-coded. Two orthogonal views are shown: side view along the membrane plane (left) and view from the cytosol (right). **(E)** Illustration of the identified variants mapped onto the 3D structure of IP_3_R3 (6DQJ). Two opposing subunits are shown in side view. Unresolved sequences are indicated by dashed lines. Red dashed lines span known phosphorylation regions. Putative residues involving Ca^2+^ binding to the receptor are rendered as pink spheres. **(F)** Slices through the space-filled model of the IP_3_R3 TM region perpendicular to the 4-fold axis as seen from the cytosol. Surfaces are color-coded according to electrostatic charges calculated for the model (red: negative; blue: positive).

Whole-exome sequencing was performed on both patients, with filtering for rare damaging variants after excluding the presence of known variants of inborn errors of immunity. We identified compound heterozygote candidate variants in *ITPR3*, encoding the Inositol-1,4,5-trisphosphate receptor subtype 3 (IP_3_R3; NM_002224.4). Three distinct variants were identified across the two compound heterozygous patients. P1 harbors a *de novo* variant, not reported in any public database but recently suggested as a candidate for causing Charcot-Marie-Tooth syndrome^12^ (c.7570C>T:p.Arg2524Cys). Additionally, P1 inherited a c.5549G>A:p.Arg1850Gln substitution from one of his parents that is also present in P2 and one of his parents (referred to as RQ^+/-^)and reported with an allelic frequency of 6%. P2 inherited a second private variant from his other parent (referred to as FL^+/-^), resulting in a c.4882T>C:p.Phe1628Leu substitution. All variants were validated by Sanger sequencing (**Supplementary Fig. 1A**) and their position within the protein is depicted in **Fig. 1B**. All protein coding variants were predicted to be damaging by the Sorting Intolerant from Tolerant^17^ (SIFT) and Deleterious Annotation of Genetic Variants Using Neural Networks^18^ (DANN) algorithms (**Supplementary Table 4**). Additionally, all variants had a CADD score above the mutation significance cut-off (MSC 5.7) and are therefore predicted to have a high impact on protein structure and function (**Fig. 1C**)^19^. These results demonstrate the identification of two reportedly unrelated patients with immunodeficiency and compound heterozygosity of predicted damaging *ITPR3* variants.

To further investigate the potential impact of the identified variants, we mapped them onto the IP_3_R3 structure solved by cryo-EM^20,21^. All altered residues are highly conserved across species, with residues F1628 and R2524 also being conserved among all isoforms of IP_3_Rs (**Supplementary Fig. 1B,C**). *In vivo*, the three isoforms IP_3_R1, IP_3_R2, and IP_3_R3 can assemble as homo- or hetero-tetramers to form the functional IP_3_-regulated Ca^2+^ channel in the ER membrane. Each IP_3_R monomer is made up of the cytosolic N-terminal IP_3_-binding domain followed by the large coupling/regulatory multi-domain region, the pore-forming region comprising six transmembrane (TM) helices and a C-terminal cytosolic tail^16,20^ (**Fig. 1B, D**). The F1628L and R1850Q variants are located in one of the regulatory alpha-helical domains, for which some flexible regions remain structurally unresolved. The R1850Q variant is close to the functionally important S1832 phosphorylation site^22^, while F1628L is in close proximity to several predicted phosphorylation and ubiquitination sites^23^ (**Fig. 1E**). The *de novo* R2524C variant is located in the sixth transmembrane domain helix (S6). The S6 helices from each subunit shape the channel’s ion conduction pathway and form the constriction gate that controls ion translocation across the ER membrane. R2524 is located in the cytosolic extension of the S6 helix just beyond the gate^21^ and its positive charge is likely neutralized through the coordination with neighboring Aspartate residues^24^. According to computational modeling, replacement of the positively charged Arginine by the uncharged Cysteine abolishes the charge coordination in the TM helix S6, thereby changing the pore charge in this region (**Fig. 1F** and **Supplementary Fig. 1D**). Due to the strong conservation of all identified variants and their location in or close to predicted functionally relevant sites we hypothesize that these variants alter channel function.

### *Functional impact of* ITPR3 *variants*

To assess the functional impact of *ITRP3* variants, we investigated expression levels and Calcium flux in patient cells. First, we analyzed the influence of the variants on mRNA and protein expression levels of all IP_3_R subtypes using isoform-specific primer pairs and antibodies selectively recognizing one of the isoforms or all of them simultaneously (panIP_3_R). The antibodies used were raised against immunogens N-terminal of the identified mutations, preserving recognition capacity of mutant IP_3_R3 proteins. mRNA expression was normal in fibroblasts (**Fig. 2A**), however *ITPR3* was reduced in both patients and the parents of P2 in peripheral blood mononuclear cells (PBMCs, **Fig. 2B**). Immunoblot analysis yielded strong immunoreactive bands for IP_3_R1 and IP_3_R3, consistent with their higher prevalence in fibroblasts and immune cells compared to low levels of IP_3_R2 (**Supplementary Fig. 2A,B**). We detected reduced levels of IP_3_R3 in P1 fibroblasts (PBMCs were not available, due to HSCT) and P2 PBMCs (**Fig. 2C,D**). Parents of P2 showed intermediate expression levels (**Fig. 2D**). While expression of IP_3_R subtypes, and the impact of variants, varies across investigated cell types, these results suggest a reduction of *ITPR3* in both patients at the mRNA and protein level in PBMCs.

**Figure 2:**
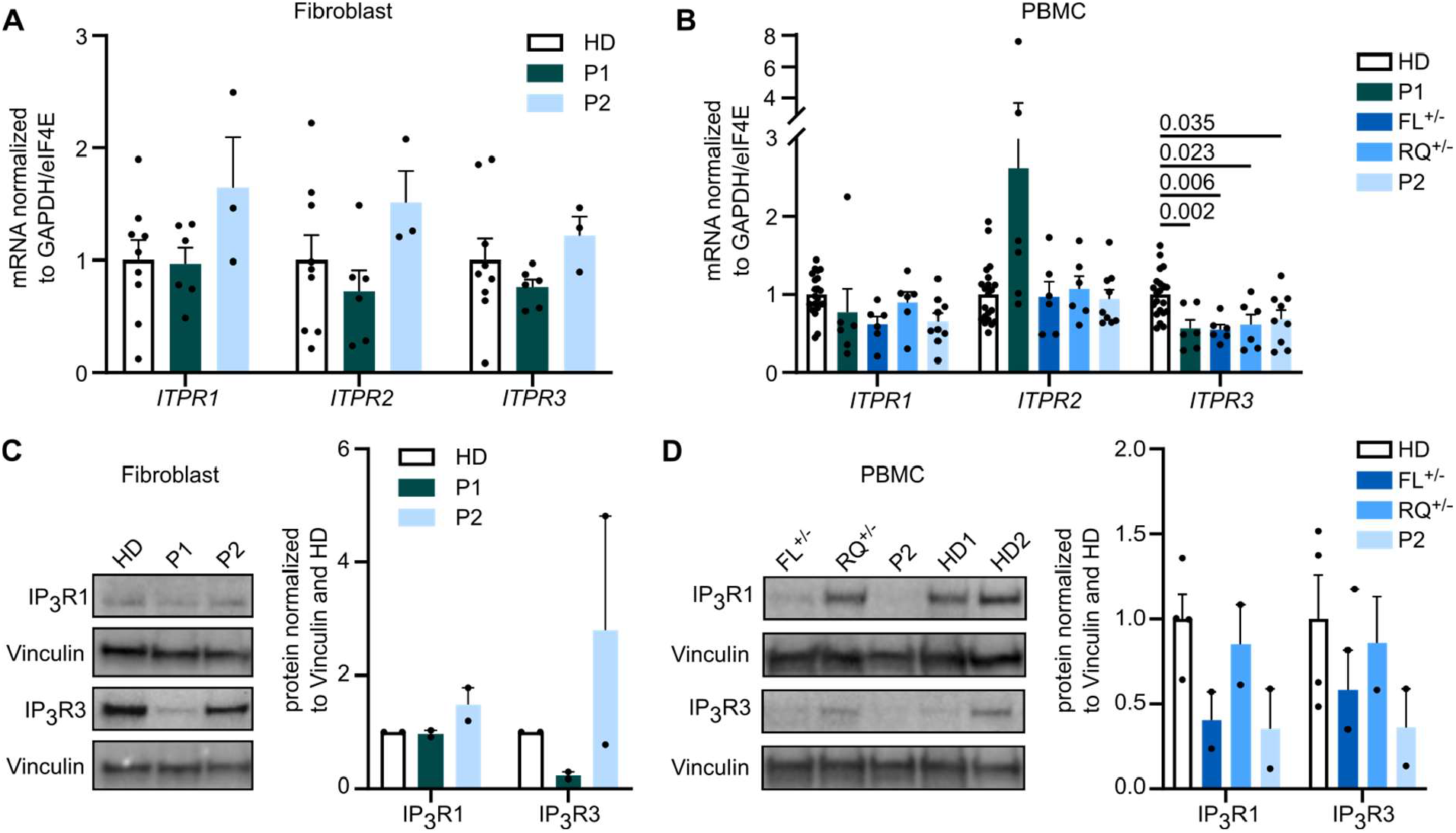
IP_3_R3 expression is reduced by variants in a cell type-dependent and variant-specific manner. mRNA expression of all three receptor subtypes was measured in primary fibroblasts and PBMCs with isoform specific primers. Dots represent the mean of duplicates for independent runs normalized to the mean values of the housekeeping genes GAPDH and eIF4E. **(A)** Expression in fibroblasts (three independent runs for HD n=3, P1 n=2, P2 n=1 biological replicates), and **(B)** PBMCs (three independent runs for HD n=7, FL^+/-^ n=2, RQ^+/-^ n=2, P2 n=3 biological replicates). **(C)** Western Blot analysis of IP_3_R2 and IP_3_R3 isoform protein expression was performed using subtype-specific antibodies. Representative blots are shown (left) with the quantification of protein expression normalized to the housekeeping gene Vinculin (right). Quantification was performed in fibroblasts (HD n=2, P1 n=2, P2 n=2) and **(D)** PBMCs (HD n=4, FL^+/-^ n=2, RQ^+/-^ n=2, P2 n=2). Values are represented as mean + SEM. One-way ANOVA with multiple comparisons.

As IP_3_Rs are crucial for Calcium dynamics and mediating IP_3_-induced Ca^2+^ flux, we sought to investigate Ca^2+^ flux in primary patient cells to test IP_3_R functionality. As pre-transplantation PBMCs were limited from P1, we used fibroblasts cultured from skin biopsies and monitored the change in [Ca^2+^]_cyt_ as a response to various stimuli using a ratiometric Ca^2+^ indicator (Fura2). We first assessed total intracellular Ca^2+^ response in fibroblasts stimulated with the Ca^2+^ ionophore Ionomycin in the presence of extracellular EGTA, a Ca^2+^-chelating agent. Both the integrated response as well as the peak amplitude were reduced in P1 (**Fig. 3A**), indicating impaired intracellular Calcium homeostasis. Likewise, P1 demonstrated reduced [Ca^2+^]_cyt_ following treatment with Thapsigargin, an inhibitor of the sarco-/endoplasmic reticulum Ca^2+^-ATPase (**Fig. 3B**), indicating a reduced steady-state ER Ca^2+^ ER store content. This could be due to an increased leakiness of Ca^2+^ through the ER membrane, thereby resulting in a lower steady-state ER Ca^2+^ concentration and thus a reduced resting concentration gradient between the ER lumen and the cytosol. Finally, we monitored IP_3_R-mediated Ca^2+^ release in fibroblasts in response to the physiological GPCR agonist Bradykinin and observed a decreased response for P1 (**Fig. 3C**). These results indicate that overall Ca^2+^ homeostasis is altered in P1, with impaired IP_3_R-mediated Ca^2+^ release. In each of these assays P2 showed a comparable response to two different healthy controls.

**Figure 3:**
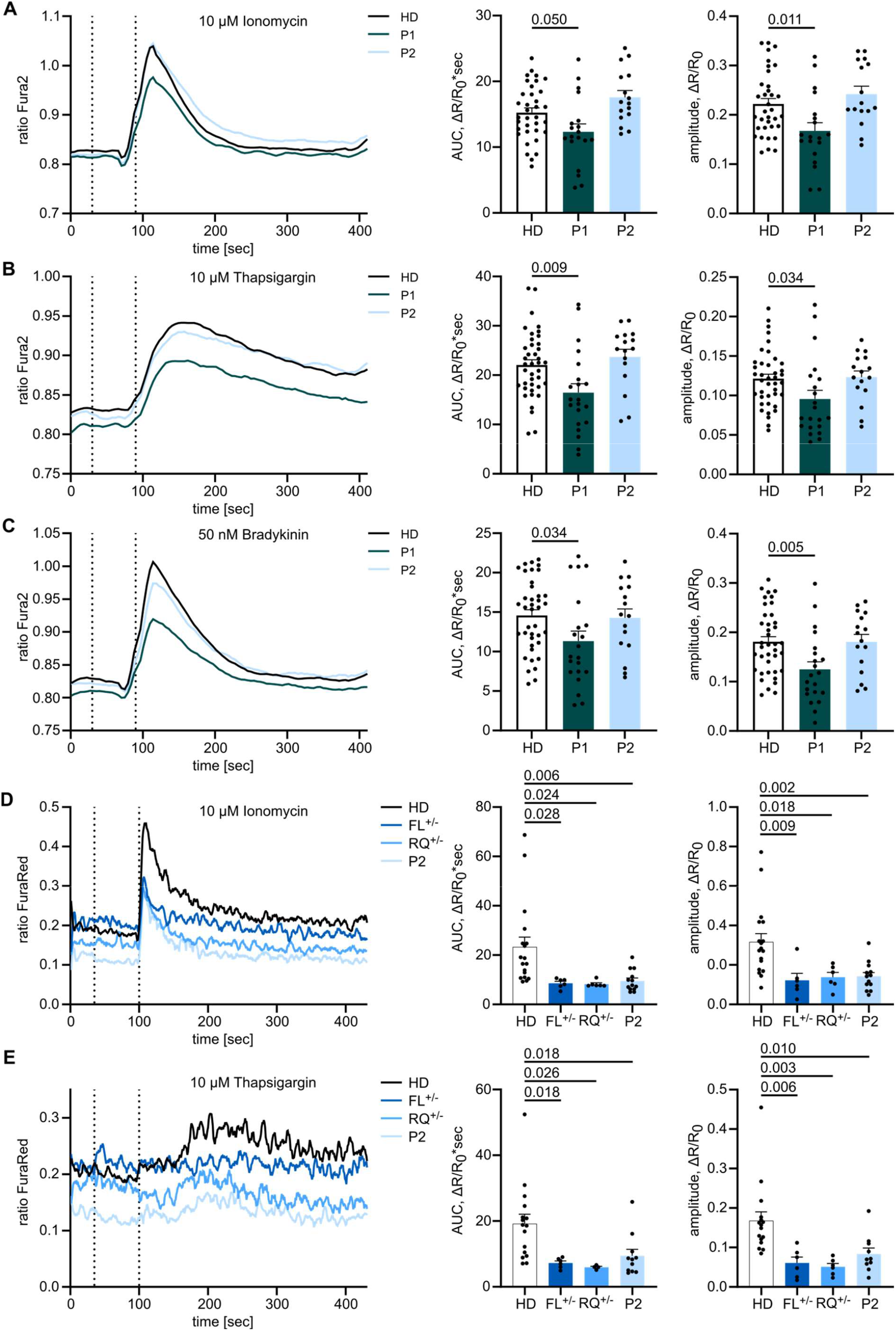
Variants in *ITPR3* impede Calcium homeostasis and Calcium signaling. Adherent fibroblasts were loaded with the ratiometric Ca^2+^ indicator Fura2/AM, and Calcium flux was monitored over time. The first dashed line indicates addition of EGTA for acquisition of background, the second dashed line indicates addition of **(A)** 10 µM Ionomycin, **(B)** 10 µM Thapsigargin or **(C)** 10 µM Bradykinin. Area under the curve (AUC) and maximal amplitude of the response were calculated respective to background values. HD n=12, P1 n=9, P2 n=4. **(D)** PBMCs were stained for CD4, loaded with the ratiometric Ca^2+^ indicator FuraRed and monitored for changes in cytosolic Ca^2+^ concentrations by flow cytometry. Mean of all experiments performed in duplicates or triplicates is depicted. Cells were treated with 10 µM Ionomycin or **(E)** 10 µM Thapsigargin. First dashed lines indicate addition of EGTA and second dashed lines indicate addition of the stimulant. HD n=7, FL^+/-^ n=3, RQ^+/-^ n=3, P2 n=5. Traces represent mean values of all experiments and response quantification is shown as mean + SEM. One-way ANOVA with multiple comparisons.

As PBMCs have a different Calcium signaling sensitivity than fibroblasts, we sought to test CD4^+^ T cells from P2 in stimulation assays (P1 was not available, due to successful HSCT). By labeling CD4^+^ T cells from different donors with different fluorochromes before combining the samples, we were able to reduce sample-to-sample variation. Cells were loaded with the ratiometric Ca^2+^ indicator FuraRed, and stimulated with Ionomycin or Thapsigargin. For both stimuli the increase in [Ca^2+^]_cyt_ was reduced in individuals of kindred 2 when compared to healthy donors (**Fig. 3D,E**). These results suggest defects in IP_3_R-mediated Ca^2+^ release in both patients, with P1 potentially the more severe (also being detected in fibroblasts), although direct comparison in PBMCs was not possible due to HSCT. Defects were also observed in the heterozygous parents of P2.

To formally test the impact of the *ITPR3* variants on Calcium signaling in a model allowing direct comparison in the absence of compensatory mechanisms, we used HEK cells that have been engineered to lack endogenous expression of all three IP_3_R subtypes^25^ (HEK-3KO) and reintroduced either wild-type (exogenous) or mutant IP_3_R3. Double-knockout of IP_3_R1 and IP_3_R2, resulting in cell lines only expressing endogenous IP_3_R3, were used as a control. We generated multiple monoclonal cell lines with transgenic expression of the mutant allele, confirmed localization to the ER by immunohistochemistry, and used Western Blot to benchmark protein expression levels against endogenous expression (**Supplementary Fig. 3A,B**). Using these knockout cell lines reconstituted with mutant or wild-type alleles, we investigated the Calcium flux induced by different concentrations of the muscarinic GPCR agonist Carbachol (CCH) and analyzed the magnitude of the response (**Supplementary Fig. 3C-E**). The inherited private F1628L variant of P2 and the inherited R1850Q variant shared by P1 and P2 resulted in reduced or absent responses under physiological protein levels, requiring more than a 40-fold over-expression to achieve similar sensitivity to the endogenous protein (**Fig. 4 A,B, Supplementary Fig. 3F,G**). The *de novo* R2524C variant of P1 showed even more severe defects, with absent Ca^2+^ mobilization across all clones (**Fig. 4C, Supplementary Fig. 3H**). Together, these results formally demonstrate IP_3_R-mediated Ca^2+^ release defects in the F1628L, R1850Q and R2524C mutants in an overexpression model, with the most striking defect being found in the mutation borne by P1, who presented with the more severe clinical manifestations of immunodeficiency.

**Figure 4:**
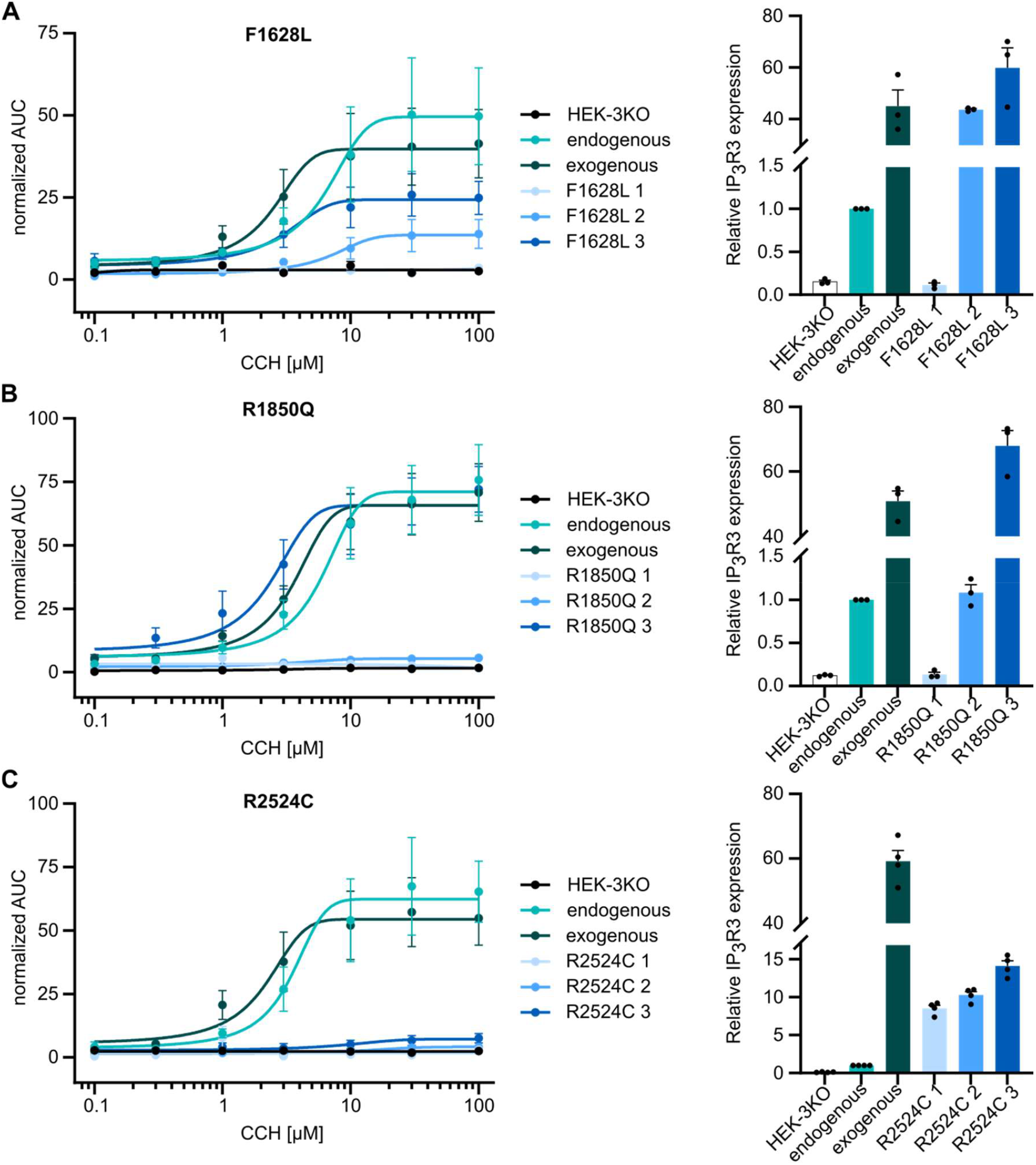
IP_3_-induced Ca^2+^ flux is reduced through F1628L, R1850Q, and R2524C mutant IP_3_R3. HEK-3KO cell lines, not expressing any IP_3_R subtype, were stably transduced with the wild-type (exogenous) or mutated versions of the IP_3_R3 receptor and stimulated with different concentrations of the GPCR agonist Carbachol in Ca^2+^ imaging buffer. Cells only expressing endogenous IP_3_R3 (endogenous) and HEK-3KO cells were used as controls. Dose-response curves of the AUC of representative cell lines stably expressing IP_3_R3 with the **(A)** F1628L (n=3), **(B)** R1850Q (n=3) or **(C)** R2524C (n=3) variant at different concentrations of Carbachol (CCh) are shown on the left. IP_3_R3 protein expression normalized to the endogenous expression was analyzed by WB and is shown on the right. Values are represented as mean ± SEM.

### *Defective immunological responses in* ITPR3 *patients*

Following our observations that Calcium homeostasis is disrupted in immune cells of our patients we investigated the downstream consequences on immune cell function. P1 was, of necessity, excluded due to successful transplantation of the hematopoietic compartment. First, we investigated the impact of variants in *ITPR3* on early downstream activation via phosphorylation of Erk and PLCγ1 in response to stimulation. Although PLCγ1 signals upstream of IP_3_Rs, it is positively regulated by Ca^2+^ and thereby amplified following IP_3_R activation^26^. Following TCR stimulation, the percentage of pPLCγ1^+^ positive cells was reduced in members of kindred 2 at different time points (**Fig. 5A,B**). Cells that responded showed comparable phosphorylation of PLCγ1 (**Fig. 5C**), indicating that the defect was in triggering the pathway. Erk activation, while highly variable among experiments, suggested defects only in P2 but not the heterozygous parents (**Fig. 5D-F**). Stimulation of the BCR gave relatively normal pErk positivity in B cells (**Fig. 5G-I**). These results suggest a functional defect of IP_3_R3 in P2 with impaired lymphocyte receptor signaling.

**Figure 5:**
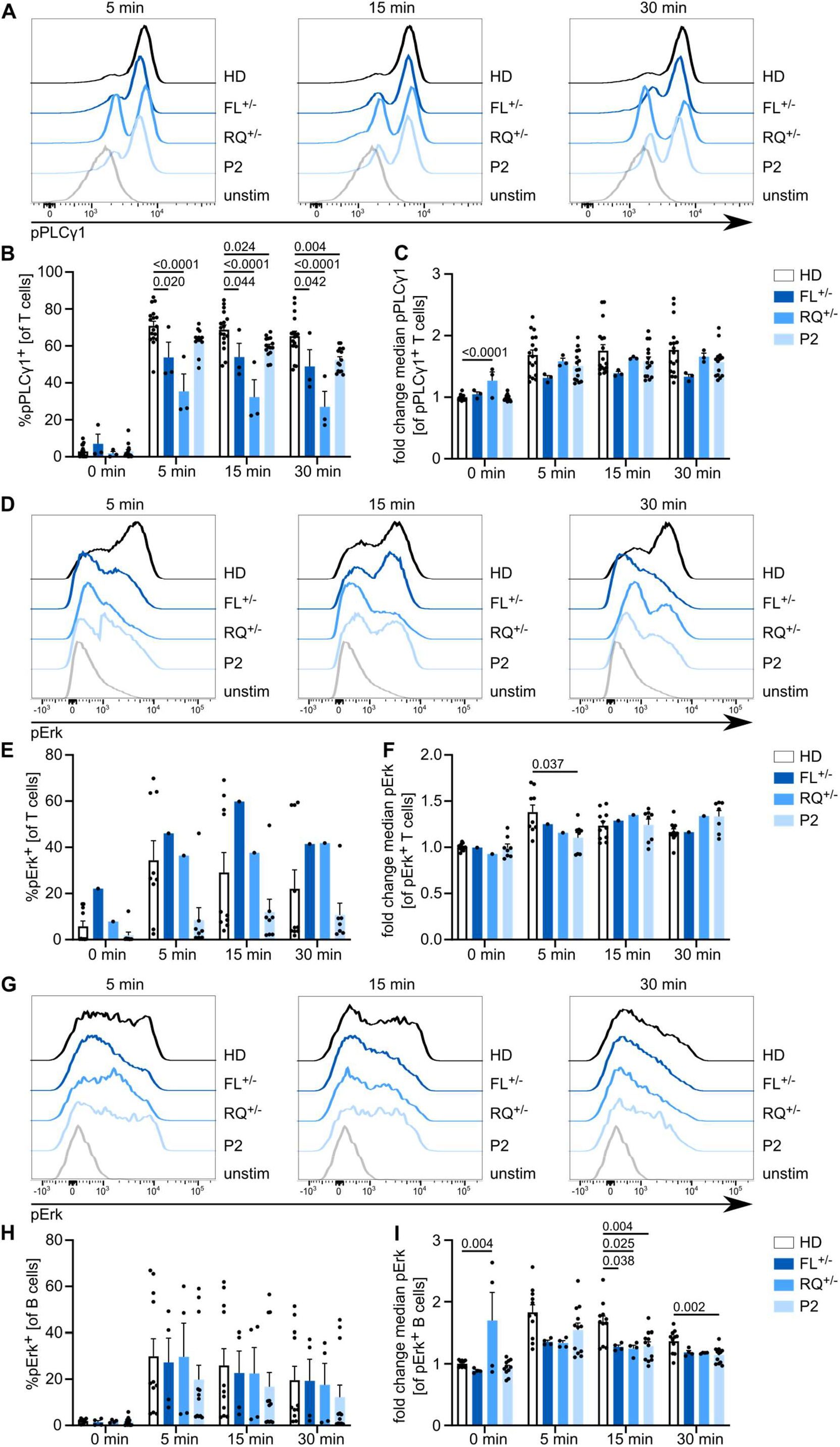
Antigen receptor downstream phosphorylation events are reduced in P2. PBMCs were stimulated with anti-CD3 and anti-CD28 or immunoglobulins for 5, 15 or 30 min. **(A)** Representative histograms illustrating PLCγ1 phosphorylation following TCR stimulation after different stimulation intervals. **(B)** Percentage of T cells with phosphorylated PLCγ1 in (un)stimulated cells. **(C)** Fold-change of median fluorescence intensity in pPLCγ1^+^ cells compared to unstimulated HD cells. HD n=10, FL^+/-^ n=2, RQ^+/-^ n=2, P2 n=6. **(D)** Representative histograms illustrating Erk phosphorylation following TCR stimulation after different stimulation intervals. **(E)** Percentage of T cells with phosphorylated Erk in (un)stimulated cells. **(F)** Fold change of median fluorescence intensity in pErk^+^ cells compared to unstimulated HD cells. HD n=7, FL^+/-^ n=1, RQ^+/-^ n=1, P2 n=3. **(G)** Representative histograms illustrating Erk phosphorylation following BCR stimulation after different stimulation intervals. **(H)** Percentage of B cells with phosphorylated Erk in (un)stimulated cells. **(I)** Fold change of median fluorescence intensity in pErk^+^ cells compared to unstimulated HD cells. HD n=7, FL^+/-^ n=2, RQ^+/-^ n=2, P2 n=5. Values are represented as mean + SEM. One-way ANOVA with multiple comparisons.

Finally, we assessed the downstream consequences of lymphocyte activation by analyzing the proliferative potential of CD4^+^ and CD8^+^ T cells in response to TCR engagement. Total cell numbers after stimulation were reduced in all individuals from kindred 2 when compared to healthy donors (**Supplementary Fig. 4A,B**). TCR-induced cell division was substantially reduced in CD4^+^ and CD8^+^ T cells of P2, with intermediate phenotypes exhibited in the heterozygous parents (**Fig. 6A,B, Supplementary Fig. 4C,D**). Interestingly, bystander activation of CD19^+^ B cells^27^ was also sharply impeded in P2 cells (**Fig. 6C, Supplementary Fig. 4E**). The reduced proliferative capacity of P2 cells was even more pronounced when stimulating PBMCs with soluble anti-CD3 and soluble anti-CD28, mimicking suboptimal stimulation conditions (**Fig. 6D-F, Supplementary Fig. 4E-G**). Taken together, our results demonstrate impaired immunological responses to TCR stimulation in P2, with weak effects observed in the heterozygous patients, consistent with the defects in Calcium flux and phosphorylation of Erk and PLCγ1.

**Figure 6:**
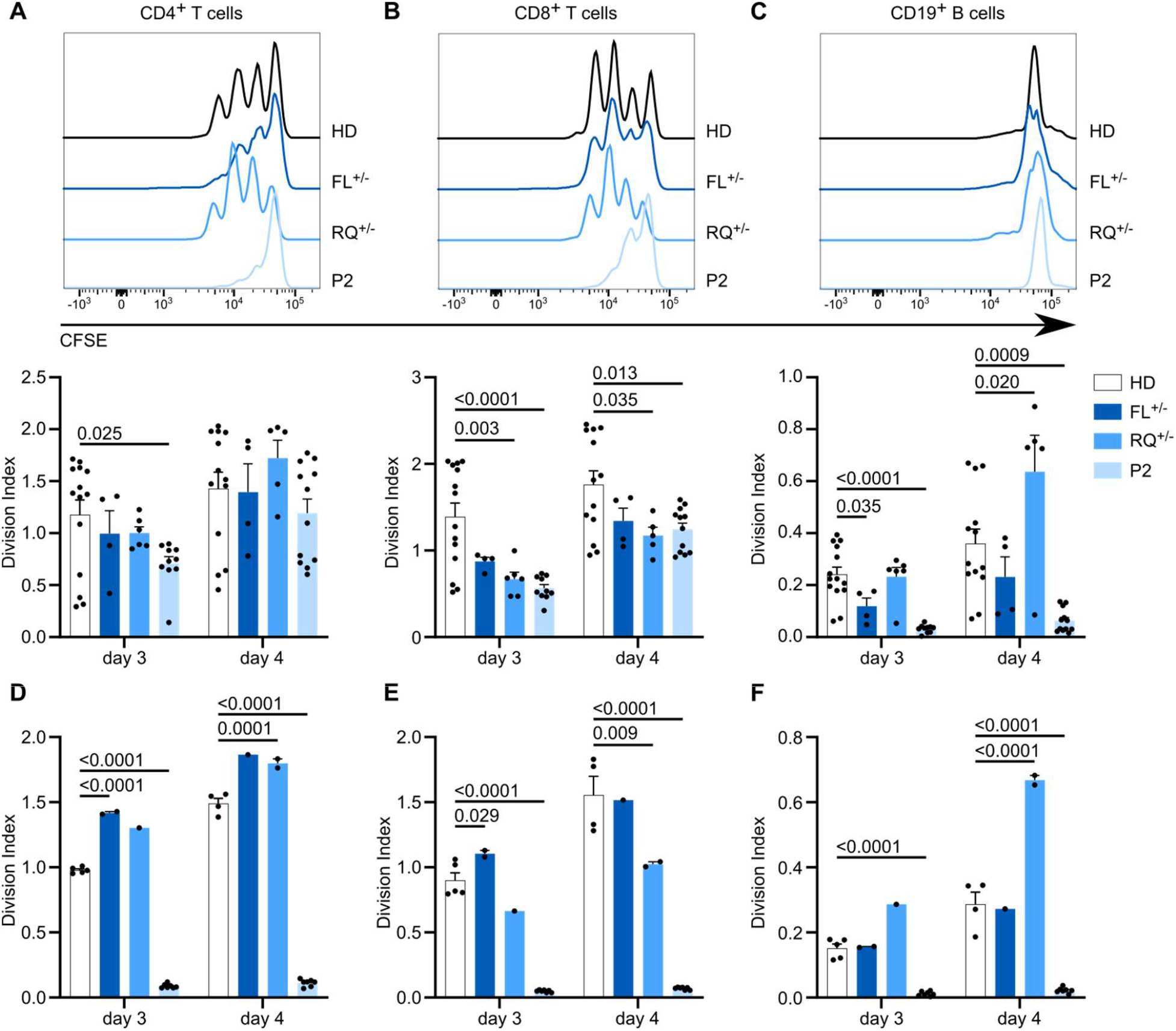
Altered cellular responses with *ITPR3* variants. PBMCs were labelled with the CFSE cell proliferation dye and stimulated with anti-CD3 and anti-CD28 for up to 4 days. The mean number of proliferations of the whole cell population, including undivided cells, was calculated for CD4^+^, CD8^+^ and CD19^+^ subsets (Division Index). Stimulation was performed in plates coated with 10 µg/mL anti-CD3 and 5 µg/mL soluble anti-CD28. Exemplary histograms are shown on top and the proliferation index for HD n=9, FL^+/-^ n=3, RQ^+/-^ n=3, P2 n=5 after 3 and 4 days shown on the bottom for CD4^+^ T cells, **(B)** CD8^+^ T cells and **(C)** B cells, reading out on bystander activation rather than direct stimulation. **(D)** Stimulation was performed with 5 µg/mL soluble anti-CD3 and 5 µg/mL soluble anti-CD28. The Division Index for HD n=2, FL^+/-^ n=1, RQ^+/-^ n=1, P2 n=3 is shown for CD4^+^ T cells, **(E)** CD8^+^ T cells and **(F)** B cells. Values are represented as mean + SEM. One-way ANOVA with multiple comparisons.

## Discussion

In this work we present two patients from unrelated families with compound heterozygous variants in *ITPR3* that present with immunodeficiency and immune dysregulation. We show that both patients exhibit altered Calcium homeostasis and responses to *in vitro* stimulation, using cell proliferation and phosphorylation of major signaling proteins as readouts. Our results suggest that the R2524C variant results in the most severe impairment of channel function, a cellular phenotype in line with the more severe combined immunodeficiency observed clinically in P1. For P2, harboring the F1628L and R1850Q variants, we demonstrate defects in all *in vitro* assays, with a profound reduction of proliferative capacity following TCR stimulation. The partially reduced sensitivity of channel activation correlates with the weaker clinical phenotype. Together with the intermediate cellular responses in the clinically healthy parents of P2, these results suggest a spectrum of escalating clinical severity with progressive decline in channel function.

For the F1628L and R1850Q variants we propose a recessive mode of inheritance based on our observations. Both variants are located in the regulatory domain of the receptor that is known for its function as signaling hub, interacting with a plethora of proteins^28,29^. We expect that each variant impairs a specific aspect of regulation, which cannot entirely be compensated for during lymphocyte activation, thus leading to signaling defects and impaired Calcium homeostasis *in vitro*. As heterozygous individuals did not show a clinical phenotype we hypothesize that lymphocytes can partially compensate for individual losses in the heterozygous state. In this model, the heterozygous loss of two regulatory partners for IP_3_R3 is sufficient to induce combined immunodeficiency in P2. An alternative explanation for the additive effect of different variants is the reported cooperative nature of channel activation, which requires all subunits of the hetero-tetramer to bind IP_3_ and undergo the necessary changes for channel gating^30^. The compound heterozygosity increases the chance of an IP_3_R containing at least one defective subunit. The *de novo* generation of the R2524C variant in an individual inheriting the R1850Q allele does not allow us to discriminate between recessive and autosomal dominant effects of this allele. However, this allele was also observed with dominant inheritance for Charcot-Marie-Tooth in Rönkkö *et al*.^12^. The absence of a reported immunological phenotype in the previously described patient suggests that sole inheritance of R2524C creates sufficient impairment to lead to Charcot-Marie-Tooth, while preserving sufficient function for normal lymphocyte activation unless coupled with an additional defective allele, as in P1. This split inheritance pattern is consistent with data demonstrating cell type- and context-dependent partial redundancies of the IP_3_R isoforms observed in mice and humans ^7,31,32^. Alternatively, the unique genetic background in the Ashkenazi Jewish patient may alter the penetrance in lymphocytes, or subclinical effects may be present which have yet to develop into a clinical phenotype.

Despite advances in the accuracy of electron microscopy techniques revealing conformational changes and interactions in specific activation states of IP_3_Rs ^20,21,24^, the effect of post-translational modifications and protein interactions remains poorly studied and new interaction partners and signaling mechanisms are continuously being discovered. Genetic variants leading to human disease provide a unique way for better understanding protein regulation and function. Our results demonstrate that the F1628L and R1850Q variants each result in reduced sensitivity to channel activation while still maintaining its gating ability. Systematic investigation of mutated receptor variants could reveal the protein interactions and/or conformational changes that are hampered and provide better insight into channel regulation. The *de novo* R2524C variant in P1, however, completely abolishes gating of a homo-tetrameric receptor. The respective conserved residue in the related Ryanodine receptor (RyR) family has indeed been shown to stabilize both the closed and open state of the channel by alternating its interaction with two other highly conserved residues^33^. In line with the prediction of a drastic change in the pore charge introduced by this variant, its counterpart in the rat IP_3_R1 protein was suggested to have a crucial function for signal transmission as mutation into different amino acids drastically reduced channel function^24,34^.

With this study we add variants in *ITPR3* as an underlying cause for disturbed Calcium signaling resulting in immunodeficiency. Although we observed a variable clinical presentation and effect on protein function depending on the location of the variants, the phenotype of the described patients resembles that of immune defects described in patients with ORAI1 and STIM1 deficiency. These patients show severely reduced or absent store-operated Ca^2+^ entry *in vitro* and present with normal lymphocyte counts but impaired T cell proliferation following *in vitro* stimulation ^4,6^. Comparable to the patients we report, they suffer from recurrent severe infections and can exhibit signs of lymphoproliferation and autoimmune disease, although the disease course is often more severe, requiring HSCT very early in life to avoid a lethal disease course ^35,36^. However, phenotypic presentation also varies greatly in STIM1 deficiency, with several STIM1 deficient patients described with late-onset or absent clinical immunodeficiency ^37,38^, and phenotypic expansion is commonly observed with increasing case recognition in primary immunodeficiency. In P1 we also observed non-immunological disease manifestations, such as mineralization defects of teeth with abnormalities of the hair and Charcot-Marie-Tooth disease, which is partly overlapping with dental enamel formation defects and myopathy - although congenital and non-progressive - in ORAI1 and STIM1 patients ^6,37,39,40^. We therefore propose that *ITPR3* should be considered as a candidate gene in patients with B and T cell defects in association with impaired Calcium signaling function, adding variants in *ITPR3* as a cause for inborn errors of immunity.

## Material and Methods

Written informed consent to conduct and publish this study was obtained from all participants and the ethics committee of UZ/KU Leuven approved the study (S52653, S58466). For details on the clinical history of the patients please contact the clinical corresponding authors.

### Whole exome sequencing and analysis

Whole exome sequencing for kindred 1 was performed using genomic DNA isolated from whole blood using the QIAmp DNA Blood Midi kit (Qiagen, Hilden, Germany) according to the manufacturer’s instructions. The sequencing library was prepared using ligation-mediated PCR and hybridizing to the SureSelect Biotinylated RNA library (Agilent Technologies, Santa Clara, CA) for enrichment and performed by BGI (Shenzhen, China) before paired-end sequencing on a HiSeq2000 (Illumina, San Diego, CA) platform. The generated FastQ files were mapped to the human genome version 19 (hg19) using the Burrows Wheeler Aligner (BWA, v0.7.5a). Duplicates were then removed using Picard MarkDuplicates and realignment performed according to the Genome Analysis Toolkit 3.1 best practices guidelines. Variants were called using the HaplotypeCaller and filtered for coding, non-synonymous variants with a high CADD score and a mean allelic frequency of <0.005 in the gnomAD database.

For kindred 2, whole exome sequencing was performed by Macrogen (Seoul, Korea). Exome capture using SureSelect Human All Exon V7 (Agilent) preceded paired-end sequencing on a NovaSeq6000 (Illumina) platform. A computational pipeline was developed using bcbio-nextgen as backend (https://github.com/bcbio/bcbio-nextgenv1.1.5-b), to process the read data and perform tasks such as quality control (QC), variant discovery, annotation and filtering. Briefly, the sequencing reads in FASTQ format were aligned to the human reference genome (GRCh37) using BWA (v0.7.17). The resulting BAM files were further processed to remove duplicate reads using biobambam (v2.0.87). Resulting variants were annotated using snpEff (v4.3.1t) for function prediction and vcfanno (v0.3.1) for adding public databases like dbsnp (v151), dbNSFP (v3.5a), ExAC, gnomAD (v r2.1), 1000 genomes (v3) and clinvar (v2019-05-13). Likely disease-causing mutations were selected and prioritized based on quality score, allele frequency, functional impact, and probable inheritance model. Sanger confirmation for individuals from both kindreds was performed by amplification of variant-specific gene products by PCR using KOD polymerase (Sigma-Aldrich, St. Louis, MO, US) and primers listed in **Supplementary Table 5**. After gel purification with the NucleoSpin Gel and PCR Clean-up (Macherey-Nagel, Düren, Germany), according to the manufacturer’s instructions, sequencing was performed by Eurofins (Ebersberg, Germany). *Variant effect prediction on protein structure*

The cryo-EM structure of IP_3_R3 with the PDB accession number 6DQJ was used to analyze the identified variants. Computational mutagenesis was performed in COOT^41^ and subsequent molecular refinements of R2524C were performed in PHENIX^42^. Calculations of electrostatic potential and all molecular visualizations were generated using UCSF Chimera^43^.

### RNA isolation and quantification

Total RNA was isolated from PBMCs and primary fibroblast cell lines generated from skin biopsies using TRIzol reagentList of Supplementary Materials (Ambion, Thermo Fisher, Waltham, MA), according to the manufacturer’s protocol. Complementary DNA was synthesized using the GoScript™ Reverse Transcription System (Promega, Madison, WI). Quantitative PCR was performed on a StepOnePlus real-time PCR system (ABI, Thermo Fisher) with Fast SYBR Green Master Mix (Applied Biosystems, Foster City, CA) supplemented with gene-specific primers (see **Supplementary Table 6**). Experiments were performed in duplicate and repeated thrice. Gene expression was normalised to the mean of two housekeeping genes before normalising to the mean expression of all healthy controls across experiments.

### Western Blot

Primary fibroblasts and PBMCs were solubilized in lysis buffer containing 20 mM Tris-HCl pH 7.5, 150 mM NaCl, 1% Triton-X, 1.5 mM MgCl_2_, 0.5 mM DTT and protease inhibitor. Protein concentrations were determined by BCA and 20 µg of protein per sample separated on a 3-8% Tris-Acetate gel. Proteins were blotted on a PVDF membrane and incubated with protein-specific primary antibodies after blocking. Primary antibodies used at a 1/1000 dilution were in-house made rabbit anti-IP_3_R1 Rbt03 antibody^44^, rabbit IP_3_R2 (Abicode C2), mouse anti-IP_3_R3 (IP3R3, BD Biosciences) and in-house made rabbit pan-IP_3_R Rbt475 antibody raised against a peptide corresponding to amino acids 127-141 of human IP_3_R1^45,46^. Vinculin (Sigma) was used as a loading control at a 1/2000 dilution. Secondary antibodies coupled to HRP were goat anti-mouse/rabbit IgG (Thermo Scientific). Membranes were developed using Pierce™ ECL (Thermo Fisher) in a Chemidoc MP Imaging system (BioRad, Hercules, CA). Quantification of protein expression was performed with ImageJ^47,48^. The fold-change in protein expression was normalized to the mean of all healthy controls within each independent experiment in Excel.

HEK-3KO cells, HEK293 cells expressing only endogenous IP_3_R3, and HEK-3KO cells stably expressing IP_3_R constructs were solubilized in RIPA lysis buffer supplemented with Halt™ protease inhibitor cocktail (Thermo Fisher). Protein concentrations were determined using D_c_ protein assay kit (BioRad) and 7.5 µg of protein per sample prepared in SDS loading buffer separated using 7.5% SDS-PAGE. Subsequently, proteins were transferred overnight to a nitrocellulose membrane (Pall Corporation, New York, NY) which was probed using the indicated primary antibodies (mouse monoclonal antibody recognizing residues 22-230 of human IP_3_R3 (BD Transduction Laboratories, San Jose, CA) and rabbit monoclonal antibody recognizing Calnexin (Cell Signaling, Danvers, MA)) and corresponding secondary antibodies. Membranes were imaged with an Odyssey infrared imaging system (LICOR Biosciences, Lincoln, NE). Resulting scans were exported to LICOR Image Studio where band intensity was calculated. In Excel, hR3 band intensity values were normalized to that of the calnexin loading control. Subsequently, these values were further normalized to that of the corresponding endogenous hIP_3_R3. Averages and statistical analysis were performed in GraphPad Prism.

### Plasmid cloning

All plasmids used in this study were based on the primary sequence of human IP_3_R3 in a pcDNA3.1 backbone. Mutagenesis to introduce base changes identified in the reported patients was performed with the primers listed in **Supplementary Table 7** that were synthesized by Integrated DNA Technologies. Suitable sequences were amplified by PCR and subcloned into pJet1.2/blunt using the CloneJET PCR Cloning kit (Thermo Fisher) according to the manufacturer’s instructions. Mutagenesis PCR was performed with outward-facing complementary primers introducing the desired base change and amplifying the full plasmid. DpnI digest in Tango Buffer (both Thermo Scientific) was performed overnight and the resulting circularized plasmids used to transform chemically competent DH5α bacteria. Following Ampicillin selection, plasmids were purified from liquid cultures of single colonies using the NucleoSpin Plasmid EasyPure kit (Macherey-Nagel) and base exchange verified by Sanger sequencing (Eurofins). Restriction-enzyme based cloning was then performed to introduce the mutated sequences into the original pcDNA3.1 construct and sequences verified again.

### Generation of HEK-3KO cell lines stably expressing IP_3_R3 mutants

HEK-3KO cells are HEK293 cells engineered though CRISPR/Cas9 for the deletion of the three endogenous IP_3_R isoforms^25^, while cells expressing endogenous IP_3_R3 are HEK293 cells modified by CRISPR/Cas9 to only express endogenous IP_3_R3. The HEK-3KO cells, cells expressing endogenous IP_3_R3, and HEK-3KO cells stably expressing exogenous IP_3_R constructs were grown at 37 °C with 5 % CO_2_ in DMEM media supplemented with 10% fetal bovine serum, 100 U/ml penicillin, 100 µg/ml streptomycin. Cell lines were maintained by subculturing as necessary and any selection required was carried out using G418 (VWR, Radnor, PA). HEK-3KO transfection was also performed similarly to previously described ^8,25^. In brief, 1×10^6^ cells were pelleted, washed once with PBS, and resuspended in a homemade transfection reagent containing ATP-disodium salt, MgCl_2_, KH_2_PO_4_, NaHCO_3_, glucose, and pH to 7.4. 2-5 µg of DNA were mixed with the resuspended cells and electroporated using the Amaxa cell nucleofector program Q-001. Cells recovered in fresh DMEM media, supplemented as listed above, for 48 h prior to passage into new 10 cm dishes containing DMEM supplemented with 1.5-2 mg/ml G418. About 7 days after start of selection individual colonies were picked by hand and transferred to 24-well plates containing fresh DMEM with G418. 10-14 days after transfection, wells that exhibited growth were expanded and screened using western blot to verify stable expression of the desired protein.

### Immunocytochemistry and Confocal Microscopy

HEK-3KO cells stably expressing exogenous IP_3_R constructs were plated on poly-d-lysine coated coverslips. When roughly 50 % confluent, cells were fixed using 4 % PFA at room temperature for 15 minutes. Subsequently, coverslips were washed with PBS and cells were blocked in 10 % Bovine Serum Albumin (BSA) in PBS-T (PBS with 0.2 % Tween20) for 1.5 h. Following blocking, cells were incubated in anti-IP_3_R3 primary antibody in BSA overnight at 4 °C. The following day, primary antibody was removed, and coverslips were washed 3 times with PBS-T for 10 minutes with gentle rocking. Subsequently, secondary antibody conjugated to AlexaFluor488 was incubated for 1 h in BSA at RT with gentle rocking. After incubation, coverslips were washed with PBS-T and mounted on slides. After allowing slides to dry, coverslips were sealed onto slides and imaged using confocal microscopy.

### Fluorescence measurement of Calcium fluxes

Fibroblasts were seeded in 96-well plates (Greiner, Kremsmünster, Austria) and assessed for [Ca^2+^]_i_ concentrations using Fura-2/AM (Eurogentec, Seraing, Belgium). Briefly, cells were loaded with the ratiometric fluorescent Ca^2+^ indicator Fura2/AM (1 µM) at RT for 30 min in a modified Krebs solution (containing 150 mM NaCl, 5.9 mM KCl, 1.2 mM MgCl_2_, 11.6 mM HEPES (pH 7.3) and 1.5 mM CaCl2). Cells were rested for 30 min at RT in the absence of Fura-2 AM to allow complete dye de-esterification before proceeding to analysis on a FlexStation 3 microplate reader (Molecular Devices, Sunnyvale, CA, USA). The indicator was alternately excited at 340 and 380 nm and emission of fluorescence at 510 nm recorded. EGTA was added after 30 sec in all conditions at a final concentration of 3 mM, to chelate Ca^2+^ ions present in the buffer, and fluorescence recorded for 60 sec prior to stimulation. Responses to stimuli, prepared in Ca^2+^-free modified Krebs solution containing 3 mM EGTA, were acquired for 6 min. Ionomycin and the irreversible SERCA-inhibitor Thapsigargin were added at a final concentration of 10 µM and Bradykinin at a final concentration of 50 nM. All traces are shown as the ratio of both emission wavelengths F_340_/F_380_.

PBMCs from different donors were first stained with different CD4 antibodies for multiplexing (all RPA-T4, eBioscience, BD Biosciences, Invitrogen). After mixing together they were loaded with the ratiometric fluorescent Ca^2+^ indicator dye FuraRed AM (Thermo Fisher) as described above. Cells were analyzed on a BD FACSCanto (BD Biosciences, Franklin Lakes, NJ, US). The ratio of emission at 450/50 nm after excitation at 405 nm and emission at 670 nm after excitation at 488 nm was exported using the Kinetics platform of the FlowJo™ software (v10.7.1, Ashland, OR). Raw data from both approaches were smoothened using a 2^nd^ order running average of 5 in GraphPad Prism (v9.0.0, San Diego, CA). A baseline value was calculated for each measurement as mean fluorescence after addition of EGTA and before addition of the stimulus. This baseline was used for calculation of the area under the curve (AUC) and the peak amplitude using GraphPad Prism with a minimum peak height of 10 % of the distance from minimum to maximum Y (v9.0.0). HEK-3KO cells and HEK-3KO cells stably expressing exogenous IP_3_R constructs were analyzed on a FlexStation 3 microplate reader (Molecular Devices) after loading with 4 µM Fura-2/AM (TEFLabs, Austin, TX) in complete DMEM. After 1 h, cells were harvested, washed, and resuspended in Ca^2+^ imaging buffer before dispensation into a black-walled flat bottom 96-well plate (Greiner). The plate was centrifuged (200xg for 2 min) and placed at 37 °C for 30 min prior to commencing the assay. Excitation was performed as described above and ^Ca2+^ imaging buffer or varying concentrations of CCh were added to induce IP_3_R-mediated Ca^2+^ release. Readings were taken every 4 sec for a total of 200 sec and data was exported from SoftMax® Pro Microplate Data Acquisition and Analysis software to Excel where the F_340_/F_380_ ratio was calculated. Ratios were normalized to the average of the first five ratio values and the AUC and peak amplitude calculated in GraphPad Prism as described above. Data was averaged from at least 3 individual plates and logistic curve fits were calculated in GraphPad Prism.

### Phosphorylation assay

Frozen PBMCs were thawed, plated and rested in complete RPMI at 37 °C prior to stimulation with 5 µg/mL aCD3 (UCHT1) and 5 µg/mL aCD28 (CD28.2, both eBioscience) or anti-human IgA/IgG/IgM (Jackson ImmunoResearch, West Grove, PA), for durations as indicated. Cells were fixed in 2 % paraformaldehyde and permeabilized with 100 % methanol, prior to staining for CD14 (M5E5, BioLegend), CD4 (RPA-T4, eBioscience), CD8 (SK1, eBioscience), CD19 (HIB19, eBioscience), phospho-Erk (Thr202/Tyr204, MILAN8R, eBioscience) and phospho-PLCγ1 (Tyr783, both Biolegend). Cells were acquired on a BD FACSCanto (BD Biosciences). After exclusion of doublets and monocytes via CD14, T cells were gated as being CD19^-^ and B cells as CD19^+^. The fold change of the median fluorescence intensity was calculated based on the average of all healthy controls within one repeat of the experiment.

### Proliferation assay

Frozen PBMCs were thawed in RPMI 1640 (Gibco™, ThermoFisher) supplemented with 10 % FBS (Tico Europe, Amstelveen, Netherlands) and 100 U/mL Penicillin/Streptomycin (Gibco™, ThermoFisher) prior to labelling with 1 µM CFSE (Invitrogen™, Carlsbad, CA) at 37 °C with regular shaking. The reaction was quenched with ice-cold FBS and cells plated after washing in complete medium containing 10 % FBS, 100 U/mL Penicillin, 25 mM HEPES and non-essential amino-acid. Cell stimulation was either performed in wells coated with 10 µg/mL anti-human CD3 (UCHT1) monoclonal antibodies and 5 µg/mL anti-human CD28 monoclonal antibodies (CD28.2, both eBioscience™, San Diego, CA) supplemented in the medium, or with both monoclonal antibodies supplemented in the medium at 5 µg/mL. Cells were stained with CD4 (RPA-T4, Invitrogen), CD8 (OKT-8, Invitrogen) and CD19 (HIB19, eBioscience) and analyzed by flow cytometry on a BD FACSCanto (BD Biosciences, Franklin Lakes, NJ, US) on day 3 and 4 after stimulation.

### Statistics

Ordinary One-way ANOVA with Dunnett’s multiple comparisons test was performed using GraphPad Prism 9.0.0 (San Diego, CA).

## Data Availability

All data will be made available upon reasonable request.

## List of Supplementary Materials

‐ Supplementary Table 1: Demographic, clinical, laboratory, technical and treatment data for investigated patients.
‐ Supplementary Table 2: Laboratory values of P1 at different relevant time points.
‐ Supplementary Table 3: Laboratory values of P2 at different relevant time points referred to in the manuscript.
‐ Supplementary Table 4: Protein-coding variants in *ITPR3* are predicted to be damaging.
‐ Supplementary Table 5: Primers for Sanger sequencing of identified variants.
‐ Supplementary Table 6: Primers used for subunit-specific qPCR analysis.
‐ Supplementary Table 7: Primers used for cloning.
‐ Supplementary Figure 1: Identified variants in *ITPR3* are highly conserved.
‐ Supplementary Figure 2: Western Blot analysis of IP_3_R2 and total IP_3_R expression is not reliably quantifiable.
‐ Supplementary Figure 3: Reconstitution of HEK-3KO cells with mutated IP_3_R3 results in stable protein expression and reveals impaired channel function.
‐ Supplementary Figure 4: Defective TCR-induced T cell proliferation in individuals from kindred 2.

## Funding and Acknowledgements

This work was supported by the VIB Grand Challenges Program, the KU Leuven C1 program, the European Union’s Horizon 2020 research and innovation programme under grant agreement No 779295 (to AL), and the Biotechnology and Biological Sciences Research Council (BBSRC) through Institute Strategic Program Grant funding BBS/E/B/000C0427 and BBS/E/B/000C0428 and the KU Leuven BOFZAP start-up grant (to SHB). GB is a recipient of a Hercules Foundation grant (AKUL/19/34) for the financing of equipment. GB, IS and DIY are partners of the FWO Scientific Research Network (CaSign W0.019.17N). IM and RS are FWO senior clinical investigator fellows. IM and RS are members of the European Reference Network for Rare Immunodeficiency, Autoinflammatory and Autoimmune Diseases (project ID No. 739543). JN, EVN and FS are FWO fellows. The authors acknowledge the important contributions of Tomas Luyten, Pier-Andrée Penttila and the KU Leuven FACS Core, and Stefanie Sente and Giorgia Bucciol for clinical trial support.

## Contributions

Patient selection, immunophenotypical characterization and ethics: IM, RS; Target identification: EVN, FS; Conceptualization: SHB, AL; Formal analysis: JN, EVN, LET; Investigation: JN, EVN, LET, FS, TK, KW, MG, MW, JB; Resources: IS, LDW, RS, IM, DIY, GB; Writing – original draft: JN, EVN, SHB, AL; Writing – review and editing: IM, RS, SHB, AL; Visualization: JN, LET; Supervision: SHB, AL; Funding acquisition: RS, SHB, AL

