## Supplementary Material for "Disrupted Ca^2+^ homeostasis and immunodeficiency in patients with functional Inositol 1,4,5-trisphosphate receptor subtype 3 defects"

**Supplementary Table 1: Demographic, clinical, laboratory, technical and treatment data for investigated patients.** TLC= total lung capacity, DLCO= diffusion capacity for carbon monoxide.

| | PRE-HSCT PATIENT 1 | PATIENT 2 |
|---|---|---|
| **GENETIC** | | |
| ITPR3 *VARIANT* | c.5549G>A; p.Arg1850Gln<br>c.7570C>T; p.Arg2524Cys | c.4882T>C; p.Phe1628Leu<br>c.5549G>A; p.Arg1850Gln |
| **CLINICAL SPECTRUM AND LABORATORY FINDINGS** | | |
| *HEMATOPOIETIC* | $T^-B^-NK^+$ CID | ITP<br>AIHA |
| *LYMPHOPROLIFERATION* | | GLILD |
| *GASTROINTESTINAL* | Enteropathy | Enteropathy<br>Chronic gastritis |
| *INFECTIONS* | Bronchopneumonia (chronic colonization with *H. influenzae*, *P. aeruginosa*, *S. aureus*)<br>Suspected mycobacterial disease (granulomatous airway infection)<br>Multiple viral infections (RSV, Molluscum contagiosum, Parainfluenza virus,…) | Bronchopneumonia (*H. influenzae, M. pneumoniae)*<br>Recurrent URTI<br>Invasive pulmonary aspergillosis<br>Gastrointestinal (*Blastocystis hominis*)<br>Primo CMV infection |
| *OTHER* | Brittle hair<br>Delayed dentition, conical teeth<br>Peripheral neuropathy | |
| **LABORATORY EXAMINATIONS** | Elevated IgG<br>Absent pneumococcal vaccine response | Low IgG<br>Low switched memory B cells<br>Absent pneumococcal vaccine response |
| **TECHNICAL EXAMINATIONS** | | |
| *CT SCAN* | Consolidation zones<br>Bronchiectasis | GLILD , aspergillus related lesions |
| *PULMONARY FUNCTION TEST* | Obstructive | Mild restrictive (TLC 83% predicted value) and decreased diffusion capacity (DLCO 73% predicted value) |
| *ABDOMINAL ULTRASOUND* | Cholecystolithiasis | Normal (post splenectomy) |
| *TRANSTHORACAL ECHOCARDIOGRAPHY* | Normal | NA |
| *BIOPSY* | | Lung: lymphoplasmocytic infiltrate<br>Stomach: chronic gastritis. Helicobacter pylori negative. |
| **IMMUNOMODULATORY TREATMENT (INDICATION)** | | |
| *CORTICOSTEROIDS* | | Prednisone (ITP, AIHA) |
| *ANTI-CD20* | | Rituximab (AIHA) |
| *IVIG/SCIG* | SCIG | IVIG |
| *OTHER* | | Cyclosporin (AIHA)<br>Cyclophosphamide (AIHA) |

**Supplementary Table 2: Laboratory values of P1 at different relevant time points.** Values out of range are italicized. ND=not-detectable. *corticosteroid treatment.

| | REFERENCE | LAST FOLLOW UP 6 YEARS POST -HSCT | REFERENCE | CONTROL BEFORE HSCT | RESPIRATORY EXACERBATION | ONSET OF LYMPHOPENIA, RSV INFECTION |
|---|---|---|---|---|---|---|
| **COMPLET** | | | | | | |
| *HEMOGLOBIN* | 13.0-16.0 g/dL | 13.3 | 11.0-13.5 g/dL<br>11.5-13.5 g/dL | *10.3* | *10.8* | *9.9* |
| *THROMBOCYTES* | 150-450x10$^9$/L | 193 | 150-450x10$^9$/L | 243 | 393 | 233 |
| *TOTAL LEUKOCYTES* | 4.5-13 x10$^9$/L | *3.98* | 5.5-15.5x10$^9$/L<br>6-17.5x10$^9$/L | *3.64* | *5.46* | *4.51* |
| *LYMPHOCYTES* | 2.0-8.0x10$^9$/L | *1.7* | 4.0-13.5x10$^9$/L<br>2.0-8.0x10$^9$/L | *1.3* | *1.9* | *0.4* |
| **IMMUNOGLOBULINS** | | | | | | |
| *IGG* | 5.76-12.65 g/L | *13.80* | 3.54-9.34 g/L<br>5.58-12.54 g/L | *19.4* | ND | *13.7* |
| *IGA* | 0.81-2.32 g/L | *<0.07* | 0.14-0.86 g/L | | ND | *2.95* |
| *IGM* | 0.30-1.59 g/L | 0.33 | 0.29-1.39 g/L | | ND | 0.41 |
| **LYMPHOCYTE COUNTS** | | | | | | |
| *B CELLS (CD19$^+$)* | 200-600/mm3 | 248 | 600-2700/mm$^3$<br>200-2100/mm$^3$ | *92* | ND | *301* |
| *SWITCHED B MEMORY (CD27$^+$IGM$^-$IGD$^-$)* | % of CD19$^+$ B cells | 2.8 | % of CD19$^+$ B cells | | ND | ND |
| *T CELLS (CD3$^+$)* | 800-3500/mm$^3$ | 1408 | 1600-6700/mm$^3$<br>900-4500/mm$^3$ | 995 | ND | *182* |
| *CD4$^+$ T CELLS* | 400-2100/mm$^3$ | 849 | 1000-4600/mm$^3$<br>500-2400/mm$^3$ | *217* | ND | ND |
| *CD8$^+$ T CELLS* | 200-1200/mm$^3$ | 466 | 400-2100/mm$^3$<br>300-1600/mm$^3$ | *774* | ND | ND |
| *NAÏVE T CELLS (CD27$^+$CD45RA$^+$)* | | | % of CD3 | | | |
| *NK CELLS* | 70-1200/mm$^3$ | 72 | 200-1200/mm$^3$<br>100-1000/mm$^3$ | | ND | ND |
| **LYMPHOCYTE STIMULATION** | | | | | | |
| *PHA* | ≥5 x10$^3$ cpm | 30.48 | ≥5 x10$^3$ cpm | | 280.71 | ND |
| *CANDIDA* | ≥5 x10$^3$ cpm | | ≥5 x10$^3$ cpm | | 8.73 | ND |
| *TETANUS TOXOID* | ≥5 x10$^3$ cpm | | ≥5 x10$^3$ cpm | | *0.96* | ND |
| **VACCINATION RESPONSE** | | | | | | |
| *HEPATITIS B SURFACE AS* | | | >=10 IU/L | | ND | *0* |
| *POLIOMYELITISVIRUS* | | | >= 1/8 | | ND | |
| *TYPE 1 AS* | | | | | | 1/4 |
| *TYPE 2 AS* | | | | | | *1/32* |
| *TYPE 3 AS* | | | | | | *1/128* |

**Supplementary Table 3: Laboratory values of P2 at different relevant time points.** Values out of range are italicized. NA=non-applicable, ND=not-detectable. *corticosteroid treatment.

| | REFERENCE | LAST FOLLOW UP | HOSPITALIZATION BRONCHO-PNEUMONIA | HOSPITALIZATION PRIMO CMV INFECTION | DIAGNOSIS CVID | START IVIG | RE-ADMISSION AIHA* | INITIAL PRESENTATION ITP |
|---|---|---|---|---|---|---|---|---|
| **COMPLETE** | | | | | | | | |
| *HEMOGLOBIN* | 12-18 g/dL | 16.9 | 17.1 | 14.6 | 16.8 | 14 | *2.6* | 15.9 |
| *THROMBOCYTES* | 150-450x$10^9$/L | 305 | 450 | 262 | 352 | 425 | 198 | *9* |
| *LEUKOCYTES* | 4-10x$10^9$/L | *14.69* | *19.6* | *17.64* | *10.32* | *13.4* | *84.8* | 6.7 |
| **IMMUNOGLOBULINS** | | | | | | | | |
| *IGG* | 7.51-15.6 g/L | 13.4 | *18.7* | 8.92 | 14 | *6.83* | *22.2* | 9.42 |
| *IGA* | 0.82-4.53 g/L | *4.89* | 3.9 | 3.10 | 3.41 | NA | 1.61 | 1.73 |
| *IGM* | 0.46-3.04 g/L | 1.64 | 0.92 | *0.43* | *0.44* | NA | 0.5 | *0.25* |
| **LYMPHOCYTE COUNTS** | | | | | | | | |
| *B CELLS ($CD19^+$)* | 82-476/µL | NA | 429 | NA | 250 | 352 | NA | NA |
| *SWITCHED B MEMORY ($CD27^+IGM^-IGD^-$)* | % of $CD19^+$ B cells | NA | NA | NA | ND | 1.1 | NA | NA |
| *T CELLS ($CD3^+$)* | 798-2823/µL | NA | 2056 | NA | *602* | 1385 | *3512* | NA |
| *$CD4^+$ T CELLS* | 455-1885/µL | NA | 517 | NA | *418* | 920 | *2149* | NA |
| *$CD8^+$ T CELLS* | 219-1124/µL | NA | *1461* | NA | *157* | NA | *1219* | NA |
| **LYMPHOCYTE STIMULATION** | | | | | | | | |
| *PHA* | ≥5 [x$10^3$] cpm | NA | NA | NA | 1640.91 | NA | NA | NA |
| *CANDIDA* | ≥5 [x$10^3$] cpm | NA | NA | NA | 191.14 | NA | NA | NA |

**Supplementary Table 4: Protein-coding variants in *ITPR3* are predicted to be damaging.** Gene-specific Mutation Significance cut-off (MSC) for the CADD score is 5.7. CADD: Combined Annotation Dependent Depletion. SIFT: Sorting Intolerant From Tolerant.

| variant | CADD | SIFT | DANN |
|---|---|---|---|
| p.Phe1628Leu | 25.8 | damaging | damaging |
| p.Arg1850Gln | 19.9 | tolerated | damaging |
| p.Arg2524Cys | 32.0 | damaging | damaging |

**Supplementary Table 5: Primers for Sanger sequencing of identified variants.**

| variant | FW primer (5' → 3') | RV primer (5' → 3') |
|---|---|---|
| p.Phe1628Leu | CAGACATCACGCTGTGTTCAGGG | CTCCGACTCCATGAGGTCTTG |
| p.Arg1850Gln | CAACATGAATGACCTGGGCAGCC | CTGGGAAGCACGATCTTGTGTG |
| p.Arg2524Cys | CTGGGTCCTCAATATCACAGCCC | GGCTCATGTCCTCACCTAGAAACC |

**Supplementary Table 6: Primers used for subunit-specific qPCR analysis.**

| | FW primer (5’ → 3’) | RV primer (5’ → 3’) |
|---|---|---|
| *ITPR1* | CTCAACAAACTGCACCACGC | AGGAGCTGGATCACATTGCC |
| *ITPR2* | TTCATCATGACCCATGCCGT | TCAGGATTAAGCTCTGCAGCTA |
| *ITPR3* | GCACCTTCATCCGGGGCTATAAG | CGGTAGATGAGGTCAAAGAGCAGG |

**Supplementary Table 7: Primers used for cloning.** Base changes to introduce the desired mutation are underlined.

| | FW primer (5’ → 3’) | RV primer (5’ → 3’) |
|---|---|---|
| *subcloning F1628L* | CAGACGATTGTGGTGCAGCTG | CTTTGGTGGTGGGGTCGACT |
| *subcloning R1850Q* | AGTCGACCCCACCACCAAAG | CACCTCCGAGTTCTCACGCTCTTC |
| *subcloning R2524C* | GGTGCCCTCAATCTGACCAACAAG | GCAACTAGAAGGCACAGTCGAGG |
| *mutagenesis F1628L* | CTGGCCTGAGCTGCTCCTCCTGGAGGGCAGTG | CACTGCCCTCCAGGAGGAGCAGCTCAGGCCAG |
| *mutagenesis R1850Q* | CAGCCTGCGCCAGGGGCACGAGGTGAG | CTCACCTCGTGCCCCTGGCGCAGGCTG |
| *mutagenesis R2524C* | CATCTTTGGGGTAATCATCGACACCTTCGCTGACCTGTGTAGTGAGAAGCAG | CTGCTTCTCACTACACAGGTCAGCGAAGGTGTCGATGATTACCCCAAAGATG |

**Supplementary Figure 1: Identified variants in *ITPR3* are highly conserved. (A)** Sequencing results for the variants identified in the two kindreds. The relevant base is marked in red and alternative bases and amino acids indicated below the reference. **(B)** Conservation of the relevant residues in *ITPR3* across species. **(C)** Conservation of the relevant residues within the $IP_3R$ family in humans. **(D)** View of the $IP_3R3$ channel pore structure in the plane of the R2524 amino acid. Illustration of the positively charged amino acid side chain of R2524 on the left (blue) and how it shields the negative charge of amino acid side chains of D2522 and D2518 (red). Location of the side chains in the case of the C2524 variant with its neutral side chain (cyan).

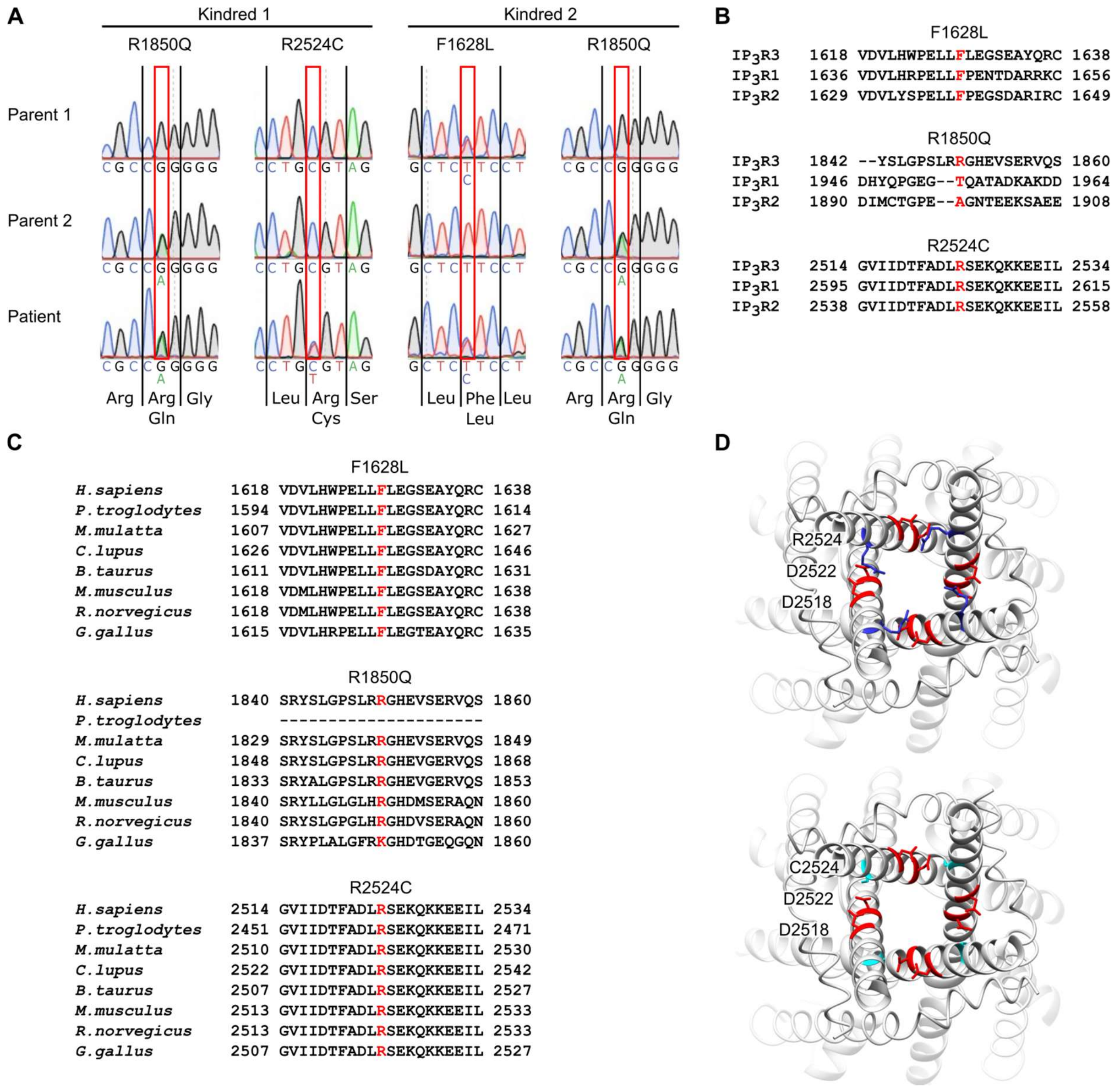
A
Kindred 1
Kindred 2
R1850Q
R2524C
F1628L
R1850Q
Parent 1
Parent 2
Patient
Arg Arg Gly
Gln
Leu Arg Ser
Cys
Leu Phe Leu
Leu
Arg Arg Gly
Gln
B
F1628L
IP3R3 1618 VDVLHWPELLFLEGSEAYQRC 1638
IP3R1 1636 VDVLHRPELLFPENTDARRKC 1656
IP3R2 1629 VDVLYSPELLFPEGSDARIRC 1649
R1850Q
IP3R3 1842 --YSLGPSLRRGHEVSERVQS 1860
IP3R1 1946 DHYQPGEG--TQATADKAKDD 1964
IP3R2 1890 DIMCTGPE--AGNTEEKSAEE 1908
R2524C
IP3R3 2514 GVIIDTFADLRSEKQKKEEIL 2534
IP3R1 2595 GVIIDTFADLRSEKQKKEEIL 2615
IP3R2 2538 GVIIDTFADLRSEKQKKEEIL 2558
C
F1628L
H.sapiens 1618 VDVLHWPELLFLEGSEAYQRC 1638
P.troglodytes 1594 VDVLHWPELLFLEGSEAYQRC 1614
M.mulatta 1607 VDVLHWPELLFLEGSEAYQRC 1627
C.lupus 1626 VDVLHWPELLFLEGSEAYQRC 1646
B.taurus 1611 VDVLHWPELLFLEGSDAYQRC 1631
M.musculus 1618 VDMLHWPELLFLEGSEAYQRC 1638
R.norvegicus 1618 VDMLHWPELLFLEGSEAYQRC 1638
G.gallus 1615 VDVLHRPELLFLEGTEAYQRC 1635
R1850Q
H.sapiens 1840 SRYSLGPSLRRGHEVSERVQS 1860
P.troglodytes ---------------------
M.mulatta 1829 SRYSLGPSLRRGHEVSERVQS 1849
C.lupus 1848 SRYSLGPSLRRGHEVGERVQS 1868
B.taurus 1833 SRYALGPSLRRGHEVGERVQS 1853
M.musculus 1840 SRYLLGLGLHRGHDMSERAQN 1860
R.norvegicus 1840 SRYSLGPGLHRGHDVSERAQN 1860
G.gallus 1837 SRYPLALGFRKGHDTGEQGQN 1860
R2524C
H.sapiens 2514 GVIIDTFADLRSEKQKKEEIL 2534
P.troglodytes 2451 GVIIDTFADLRSEKQKKEEIL 2471
M.mulatta 2510 GVIIDTFADLRSEKQKKEEIL 2530
C.lupus 2522 GVIIDTFADLRSEKQKKEEIL 2542
B.taurus 2507 GVIIDTFADLRSEKQKKEEIL 2527
M.musculus 2513 GVIIDTFADLRSEKQKKEEIL 2533
R.norvegicus 2513 GVIIDTFADLRSEKQKKEEIL 2533
G.gallus 2507 GVIIDTFADLRSEKQKKEEIL 2527
D
R2524
D2522
D2518
C2524
D2522
D2518

**Supplementary Figure 2: Western Blot analysis of $IP_3R2$ and total $IP_3R$ expression is not reliably quantifiable. (A)** Western Blot analysis of $IP_3R2$ protein expression was performed using a subtype-specific antibody as well as a non-specific antibody (pan-$IP_3R$). Representative blots are shown for fibroblasts and **(B)** PBMCs indicating low $IP_3R2$ expression.

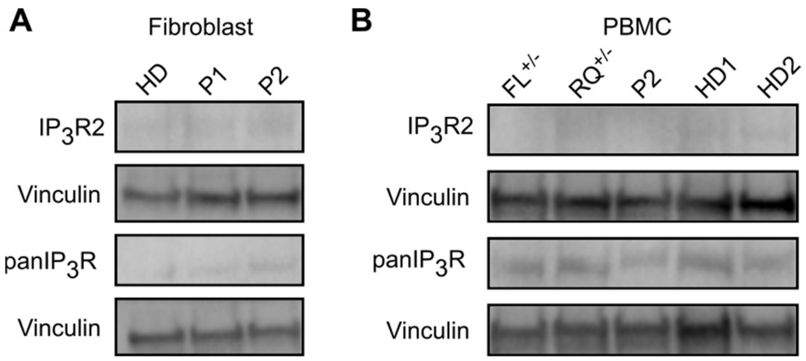
A
Fibroblast
HD
P1
P2
IP3R2
Vinculin
panIP3R
Vinculin
B
PBMC
FL+/-
RQ+/-
P2
HD1
HD2
IP3R2
Vinculin
panIP3R
Vinculin

**Supplementary Figure 3: Reconstitution of HEK-3KO cells with mutated $IP_3R3$ results in stable protein expression and reveals impaired channel function. (A)** Immunocytochemistry for $IP_3R3$ (green) was performed to validate physiological localization in HEK-3KO cell lines expressing either exogenous wild-type $IP_3R3$ (top) or mutant $IP_3R3$ (bottom). Representative images are shown. Red scale bars, 5 µm. **(B)** Exogenous wild-type $IP_3R3$ and mutant $IP_3R3$ stable cell lines were western blotted alongside HEK293-3KO and HEK293 cell lines modified to express only endogenous wild-type $IP_3R3$ (endogenous $IP_3R3$). Representative blots are shown. **(C)** HEK-3KO cell lines stably transduced with the wild-type (exogenous) or mutated versions of the $IP_3R3$ receptor were stimulated with different concentrations of the GPCR agonist Carbachol in $Ca^{2+}$ imaging buffer. Cells only expressing endogenous $IP_3R3$ (endogenous) and HEK-3KO cells were used as controls. Representative traces of one well stimulated with 100 µM CCH are shown for F1628L, **(D)** R1850Q or **(E)** R2524C as 340/380 ratio normalized to the first 5 values of the measurement. **(F)** Dose-response curves of the peak amplitude of representative cell lines stably expressing $IP_3R3$ with the F1628L (n=3), **(G)** R1850Q (n=3) or **(H)** R2524C (n=3) variant at different concentrations of Carbachol (CCH) are shown as mean ± SEM.

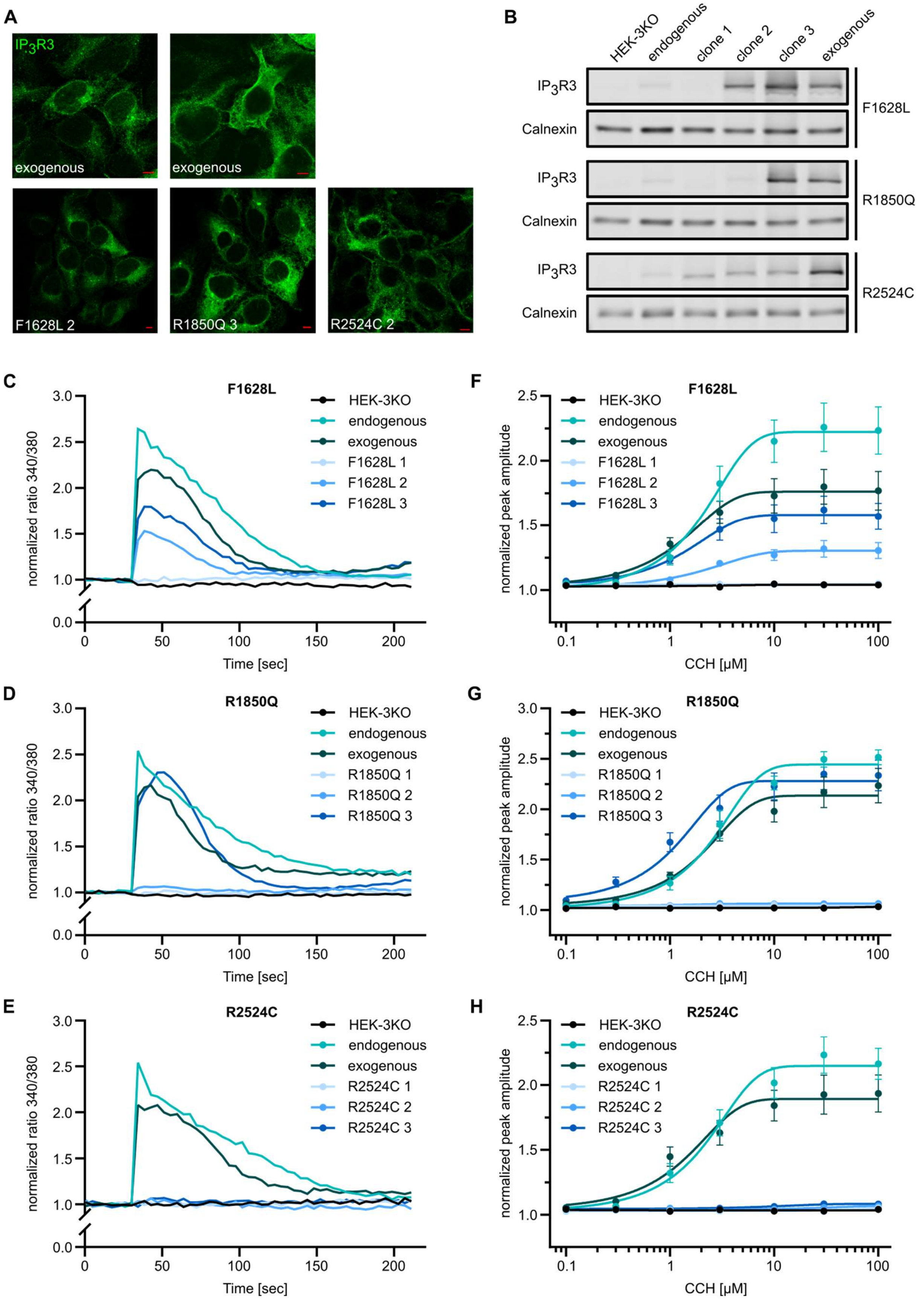

A
IP3R3
exogenous
exogenous
F1628L 2
R1850Q 3
R2524C 2
B
HEK-3KO
endogenous
clone 1
clone 2
clone 3
exogenous
IP3R3
Calnexin
F1628L
IP3R3
Calnexin
R1850Q
IP3R3
Calnexin
R2524C
C
F1628L
HEK-3KO
endogenous
exogenous
F1628L 1
F1628L 2
F1628L 3
normalized ratio 340/380
Time [sec]
D
R1850Q
HEK-3KO
endogenous
exogenous
R1850Q 1
R1850Q 2
R1850Q 3
normalized ratio 340/380
Time [sec]
E
R2524C
HEK-3KO
endogenous
exogenous
R2524C 1
R2524C 2
R2524C 3
normalized ratio 340/380
Time [sec]
F
F1628L
HEK-3KO
endogenous
exogenous
F1628L 1
F1628L 2
F1628L 3
normalized peak amplitude
CCH [μM]
G
R1850Q
HEK-3KO
endogenous
exogenous
R1850Q 1
R1850Q 2
R1850Q 3
normalized peak amplitude
CCH [μM]
H
R2524C
HEK-3KO
endogenous
exogenous
R2524C 1
R2524C 2
R2524C 3
normalized peak amplitude
CCH [μM]

**Supplementary Figure 4: Defective TCR-induced T cell proliferation in individuals from kindred 2.** PBMCs were labelled with the CFSE cell proliferation dye and stimulated with 10 μg/mL plate-bound anti-CD3 and 5 μg/mL soluble anti-CD28 for 3 and 4 days. **(A)** Total cell numbers in unstimulated or stimulated wells after 3 and **(B)** 4 days. The mean number of proliferations of cells that divided at least once (Proliferation Index), was calculated for **(C)** $CD4^+$, **(D)** $CD8^+$ and **(E)** $CD19^+$ subsets. HD n=9, $FL^{+/-}$ n=3, $RQ^{+/-}$ n=3, P2 n=5. **(F)** Stimulation was performed with 5 μg/mL soluble anti-CD3 and 5 μg/mL soluble anti-CD28. The Proliferation Index for HD n=2, $FL^{+/-}$ n=1, $RQ^{+/-}$ n=1, P2 n=3 is shown for $CD4^+$ T cells, **(G)** $CD8^+$ T cells and **(H)** B cells Values are represented as mean + SEM. One-way ANOVA with multiple comparisons.

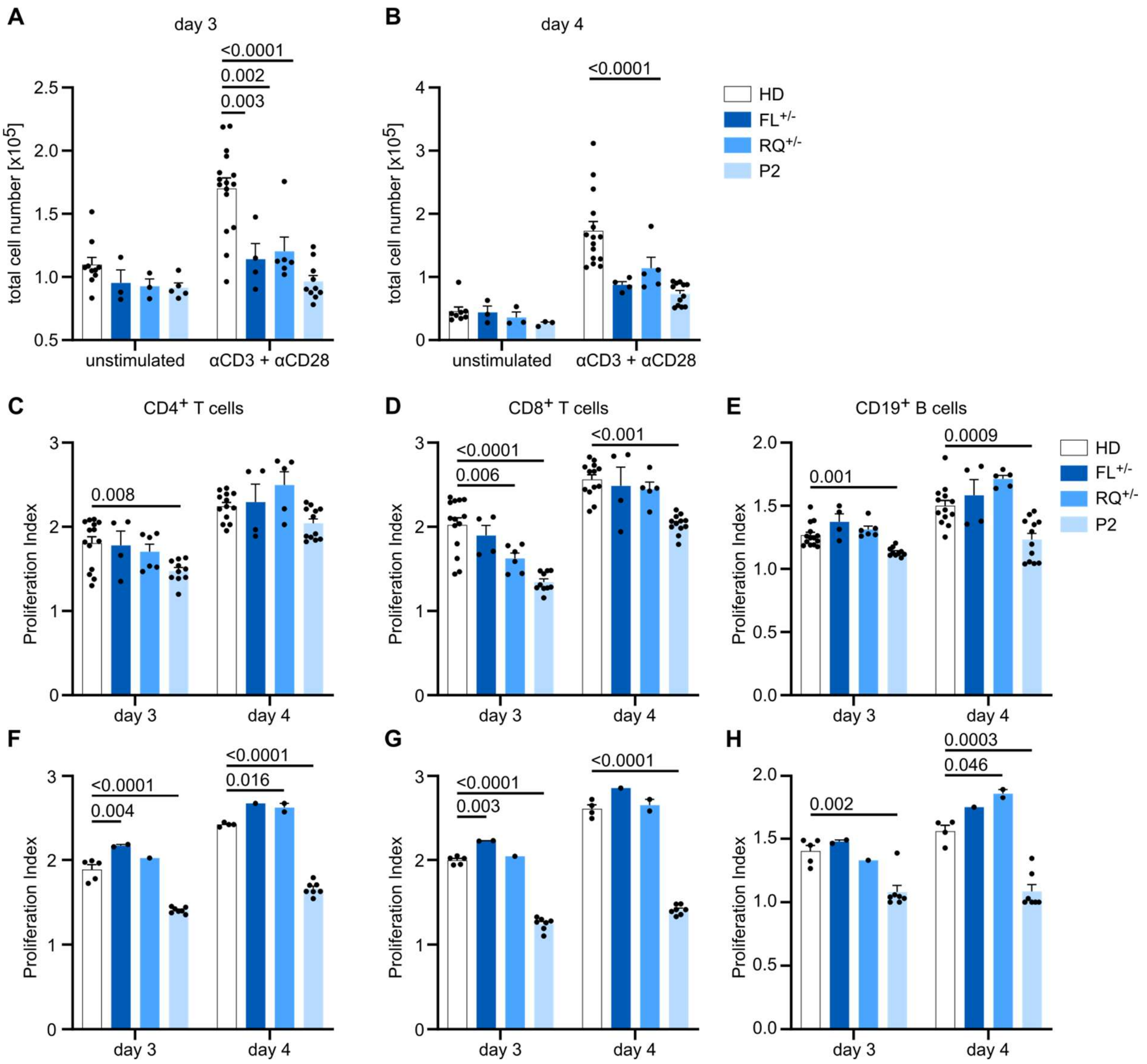
A
day 3
<0.0001
0.002
0.003
total cell number [x10^5]
2.5
2.0
1.5
1.0
0.5
unstimulated
αCD3 + αCD28
B
day 4
<0.0001
total cell number [x10^5]
4
3
2
1
0
unstimulated
αCD3 + αCD28
HD
FL+/-
RQ+/-
P2
C
CD4+ T cells
0.008
Proliferation Index
day 3
day 4
D
CD8+ T cells
<0.0001
0.006
<0.001
Proliferation Index
day 3
day 4
E
CD19+ B cells
0.0009
0.001
Proliferation Index
day 3
day 4
HD
FL+/-
RQ+/-
P2
F
<0.0001
0.004
<0.0001
0.016
Proliferation Index
day 3
day 4
G
<0.0001
0.003
<0.0001
Proliferation Index
day 3
day 4
H
0.0003
0.046
0.002
Proliferation Index
day 3
day 4
